# Unique Capabilities of Genome Sequencing for Rare Disease Diagnosis

**DOI:** 10.1101/2023.08.08.23293829

**Authors:** Monica H Wojcik, Gabrielle Lemire, Maha S Zaki, Mariel Wissman, Wathone Win, Sue White, Ben Weisburd, Leigh B Waddell, Jeffrey M Verboon, Grace E. VanNoy, Ana Töpf, Tiong Yang Tan, Volker Straub, Sarah L Stenton, Hana Snow, Moriel Singer-Berk, Josh Silver, Shirlee Shril, Eleanor G Seaby, Ronen Schneider, Vijay G Sankaran, Alba Sanchis-Juan, Kathryn A Russell, Karit Reinson, Gianina Ravenscroft, Eric A Pierce, Emily M Place, Sander Pajusalu, Lynn Pais, Katrin Õunap, Ikeoluwa Osei-Owusu, Volkan Okur, Kaisa Teele Oja, Melanie O’Leary, Emily O’Heir, Chantal Morel, Rhett G Marchant, Brian E Mangilog, Jill A Madden, Daniel MacArthur, Alysia Lovgren, Jordan P Lerner-Ellis, Jasmine Lin, Nigel Laing, Friedhelm Hildebrandt, Emily Groopman, Julia Goodrich, Joseph G Gleeson, Roula Ghaoui, Casie A Genetti, Hanna T Gazda, Vijay S. Ganesh, Mythily Ganapathy, Lyndon Gallacher, Jack Fu, Emily Evangelista, Eleina England, Sandra Donkervoort, Stephanie DiTroia, Sandra T Cooper, Wendy K Chung, John Christodoulou, Katherine R Chao, Liam D Cato, Kinga M Bujakowska, Samantha J Bryen, Harrison Brand, Carsten Bonnemann, Alan H Beggs, Samantha M Baxter, Pankaj B Agrawal, Michael Talkowski, Chrissy Austin-Tse, Heidi L Rehm, Anne O’Donnell-Luria

**Affiliations:** Divisions of Newborn Medicine and Genetics and Genomics, Department of Pediatrics, Boston Children’s Hospital, Harvard Medical School, Boston, Massachusetts, USA; Manton Center for Orphan Disease Research, Division of Genetics and Genomics, Department of Pediatrics, Boston Children’s Hospital, Harvard Medical School, Boston, Massachusetts, USA; Broad Center for Mendelian Genomics, Broad Institute of MIT and Harvard, Cambridge, Massachusetts, USA; Clinical Genetics Department, Human Genetics and Genome Research Instiute, National Research Centre, Cairo, Egypt; Division of Hematology and Oncology, Boston Children’s Hospital, Harvard Medical School, Boston, Massachusetts, USA; Department of Pediatric Oncology, Dana-Farber Cancer Institute, Harvard Medical School, Boston, Massachusetts, USA; Broad Institute of MIT and Harvard, Cambridge, Massachusetts, USA; Victorian Clinical Genetics Service, Murdoch Children’s Research Institute, Parkville, Australia; Department of Pediatrics, University of Melbourne, Melbourne, Australia; Kids Neuroscience Centre, Kids Research, Children’s Hospital at Westmead, Westmead, New South Wales, Australia; Discipline of Child and Adolescent Health, Faculty of Medicine and Health, The University of Sydney, Westmead, New South Wales, Australia; John Walton Muscular Dystrophy Research Centre, Clinical and Translational Research Institute, Newcastle University and NHS Trust, Newcastle upon Tyne, United Kingdom; Victorian Clinical Genetics Services, Melbourne, Australia; Murdoch Children’s Research Institute, Melbourne, Australia; Fred A. Litwin Family Centre in Genetic Medicine, University Health Network, Toronto, Ontario, Canada; Department of Molecular Genetics, University of Toronto, Toronto, Ontario, Canada; Department of Pediatrics, Boston Children’s Hospital, Harvard Medical School, Boston, Massachusetts, USA; Harvard Stem Cell Institute, Cambridge, Massachusetts, USA; Center for Genomic Medicine, Massachusetts General Hospital, Boston, Massachusetts, USA; Medical and Population Genetics, Broad Institute of MIT and Harvard, Cambridge, Massachusetts, USA; Department of Neurology, Harvard Medical School, Boston, Massachusetts, USA; Department of Clinical Genetics, Genetics and Personalized Medicine Clinic, Tartu University Hospital, Tartu, Estonia; Department of Clinical Genetics, Institute of Clinical Medicine, University of Tartu, Tartu, Estonia; Harry Perkins Institute of Medical Research and Centre for Medical Research, University of Western Australia; Ocular Genomics Institute, Department of Ophthalmology, Massachusetts Eye and Ear, Harvard Medical School, Boston, Massachusetts, USA; Molecular Diagnostics, New York Genome Center, New York, USA; Faculty of Medicine, University of Toronto, Toronto, Ontario, Canada; Functional Neuromics, Children’s Medical Research Institute, Westmead, New South Wales, Australia; Pathology and Laboratory Medicine, Mount Sinai Hospital, Sinai Health, Toronto, Ontario, Canada; Department of Laboratory Medicine and Pathobiology, University of Toronto, Toronto, Ontario, Canada; Lunenfeld Tanenbaum Research Institute, Mount Sinai Hospital, Sinai Health, Toronto, Ontario, Canada; Department of Neurosciences, Rady Children’s Institute for Genomic Medicine, University of California, San Diego, La Jolla, California, USA; Department of Neurology, Central Adelaide Local Health Network/Royal Adelaide Hospital, Adelaide, SA, Australia; Adelaide Medical School, The University of Adelaide, Adelaide, SA, Australia; Department of Genetics & Molecular Pathology, SA Pathology, Adelaide, SA, Australia; Department of Neurology, Brigham and Women’s Hospital, Boston, Massachusetts; Department of Pathology & Cell Biology, Columbia University Irving Medical Center, New York, USA; Victorian Clinical Genetics Services, Murdoch Children’s Research Institute and Department of Paediatrics, University of Melbourne, Melbourne, Victoria, Australia; Neuromuscular and Neurogenetic Disorders of Childhood Section, National Institute of Neurological Disorders and Stroke, National Institutes of Health, Bethesda, Maryland, USA; Brain and Mitochondrial Research Group, Murdoch Children’s Research Institute and Department of Paediatrics, University of Melbourne, Melbourne, Victoria, Australia; Division of Neonatology, Department of Pediatrics, University of Miami School of Medicine and Holtz Children’s Hospital, Jackson Health System, Miami, FL; Stanley Center for Psychiatric Research, Broad Institute of MIT and Harvard, Cambridge, Massachusetts, USA

**Author notes:** Corresponding author: Monica H Wojcik, MD, MPH, 300 Longwood Ave BCH 3036, Boston, MA, 02115; Anne O’Donnell-Luria,;, 300 Longwood Ave, Boston, MA, 02115.

## Abstract

**Background:** Causal variants underlying rare disorders may remain elusive even after expansive gene panels or exome sequencing (ES). Clinicians and researchers may then turn to genome sequencing (GS), though the added value of this technique and its optimal use remain poorly defined. We therefore investigated the advantages of GS within a phenotypically diverse cohort.

**Methods:** GS was performed for 744 individuals with rare disease who were genetically undiagnosed. Analysis included review of single nucleotide, indel, structural, and mitochondrial variants.

**Results:** We successfully solved 218/744 (29.3%) cases using GS, with most solves involving established disease genes (157/218, 72.0%). Of all solved cases, 148 (67.9%) had previously had non-diagnostic ES. We systematically evaluated the 218 causal variants for features requiring GS to identify and 61/218 (28.0%) met these criteria, representing 8.2% of the entire cohort. These included small structural variants (13), copy neutral inversions and complex rearrangements (8), tandem repeat expansions (6), deep intronic variants (15), and coding variants that may be more easily found using GS related to uniformity of coverage (19).

**Conclusion:** We describe the diagnostic yield of GS in a large and diverse cohort, illustrating several types of pathogenic variation eluding ES or other techniques. Our results reveal a higher diagnostic yield of GS, supporting the utility of a genome-first approach, with consideration of GS as a secondary or tertiary test when higher-resolution structural variant analysis is needed or there is a strong clinical suspicion for a condition and prior targeted genetic testing has been negative.

## Introduction

Massively-parallel (“next generation”) sequencing has revolutionized clinical medicine by uncovering the causal variants underlying rare conditions, particularly via large gene panels or exome sequencing (ES).^1^ Even if a clinical diagnosis has been made, identifying the molecular diagnosis – the underlying genomic change responsible for disease – can provide new clinical insight and support individualized therapies^2^ and familial testing and counseling. However, many causal variants, including those uniquely amenable to precision therapies,^3^ remain elusive even after ES or gene panel sequencing. Thus, a majority of rare disease patients remain molecularly- undiagnosed,^4,5^ and clinicians may thus turn to genome sequencing (GS). Although more costly than ES, which only evaluates the ∼2% of the genome that is protein-coding, GS has many potential advantages, including the ability to detect variants that ES cannot easily detect, such as certain structural variants (SVs, encompassing a broader range of copy number gains and losses in addition to copy-neutral inversions, retroviral insertions and other, more complex events), tandem repeat expansions (TREs), and deep intronic variants. Moreover, the lack of an exon-capture step in GS leads to improved uniformity of sequence coverage, increasing detection sensitivity for single nucleotide variants (SNVs) and insertions/deletions (indels) in certain coding regions compared to ES.^6–10^ However, with the enhanced technical sensitivity of GS comes a higher analytic burden due to the millions of non-coding or structural variants identified. To date, the few publications that have compared the relative yield of GS versus ES have found that higher detection sensitivity of GS is associated with only a modest increase in diagnostic yield.^11–14^ Thus, the incremental benefit of GS remains unclear.

Through the Center for Mendelian Genomics (CMG) and the Rare Genomes Project (RGP) at the Broad Institute of MIT and Harvard, we have sequenced and analyzed over 8,000 families with rare, suspected monogenic disease, employing a variety of analytic techniques to identify pathogenic variants responsible for a variety of phenotypes.^15^ We therefore evaluated the diagnostic yield of GS for rare disease diagnosis within this cohort, focusing on features enabling successful diagnosis via GS, particularly where ES or other methods had been unsuccessful.

## Methods

The Broad Institute CMG and RGP are research studies aimed at genetic diagnosis and discovery for people with suspected rare Mendelian disorders. The Broad CMG was established in 2016 as part of an initiative funded by the National Institutes of Health to identify novel disease genes underlying Mendelian disorders using ES and GS.^15–17^ Families sequenced through the Broad CMG are enrolled by collaborating investigators in research studies under a local institutional review board (IRB)-approved protocol that includes a provision for data sharing. RGP was launched in 2017 as a patient-centered initiative to directly recruit individuals and families with suspected Mendelian disorders throughout the United States for sequencing and analysis via the Broad CMG. Potential participants self-refer through the study website. Both studies have been approved by the Mass General Brigham IRB.

Over a five-year period, from April 2016 to April 2021, 744 families underwent GS via the Broad CMG (354 families) or RGP (390 families) following an unrevealing prior diagnostic evaluation.

### Case selection

Individuals or families were selected for GS after case review by at least two members of CMG/RGP team (genetic counselors, clinical geneticists or other relevant subspecialists). Cases approved for GS were thought to have a high likelihood of an underlying genetic disorder in addition to appropriate, non-diagnostic prior testing justifying the need for GS, such as ES or targeted testing of specific disease genes. The RGP study also includes families who are unable to access genetic testing due to lack of insurance coverage or other barriers to care and, therefore, may have not had any prior genetic testing.

### Sequencing and Analysis

GS and data processing were performed by the Genomics Platform at the Broad Institute. Our approaches to data analysis continue to evolve as we develop innovative approaches to derive the maximum diagnostic yield. Additionally, our Broad CMG collaborators use multiple “homegrown” approaches to data analysis based on their experience and the genetic architecture of the various phenotypes. Generally, we analyze our ES/GS data under the assumptions that the affected individual/family has a severe, rare Mendelian condition, and our analysis filters reflect these assumptions (Table S1). Our “first pass” analysis for SNV and indels includes evaluation for both *de novo* dominant or recessive conditions. For most families, we also perform a search with a list of priority genes based on phenotype and incorporating literature reports, Online Mendelian Inheritance in Man (OMIM)^18^, and internal communication with researchers/collaborators and relax our filtering strategy when using these lists. Additional sequencing methods and analytic tools are further described in the Supplement.

### Variant interpretation

Candidate variants (SNVs, indels and SVs) in known disease-associated genes were classified according to established criteria^19–21^ (additional details in the Supplement). We considered a case solved if a pathogenic or likely pathogenic variant was found in a known disease gene that explained the phenotype or if a variant was found in a novel disease gene with moderate/strong supporting evidence for the individual’s phenotype by ClinGen criteria^22^; we considered cases likely solved when a variant was identified in a known disease gene that was classified as a variant of uncertain significance (VUS) by ACMG/AMP criteria but the multidisciplinary CMG team and referring provider, when relevant, considered the variant causative based upon supportive clinical data. Re-analysis of unsolved cases remains ongoing; the solve status reported here is as of May 1, 2023.

### Evaluation of diagnoses

We systematically evaluated all solved/likely solved cases to determine variants requiring GS for identification (Figure). These included deep intronic non-coding variants as well as coding variants such as SNVs and indels poorly covered by standard ES platforms. In addition, we evaluated all TREs and SVs to identify those that were missed on prior exome or could not reliably be found using ES data. For example, larger CNVs in well-covered regions may be identified on ES data if dedicated CNV calling pipelines are applied. Because the performance of many CNV callers on ES data decays for CNVs smaller than 3 exons, ^23–25^ any heterozygous CNV with less than 3 targets was considered to require GS as it would not be reliably detected by ES. We also considered CNVs involving noncoding regions or regions that are challenging to sequence (such as high sequence homology, exons with high GC content) as requiring GS to detect and manually reanalyzed the ES data when available with updated CNV calling to see if we could retrospectively identify the CNV. Comparisons of diagnostic yield across categories were made using the Fisher’s exact test or chi square test when appropriate.

## Results

### Diagnostic yield

Of the 744 families who underwent GS, we consider 218 (29.3%) solved or likely solved (Table S2). Characteristics of the probands undergoing GS are presented in Table 1. Most solves (157/218, 72.0%) were identified in previously known disease genes and the remainder represented novel disease gene discoveries (61/218, 32.1%), with one case involving both a novel gene and a known gene contributing to a blended phenotype. Our diagnostic yield was lowest in cases sequenced as duos (two affected siblings or a parent/proband pair, 14/77, 18.2%), compared to proband only (44/215, 20.5%), trios (136/369, 36.9%), or larger family groups (24/83, 28.9%) (*p* < 0.001). Diagnostic yield was lower in RGP-enrolled cases (102/390, 26.1%) compared to CMG collaborator-recruited cases (116/354, 32.8%), though this did not reach statistical significance. Although we allowed for cases to be considered solved by VUS that were clinically-interpreted by our multi-disciplinary team as causal, excluding any solve involving a VUS would result in a diagnostic yield of 130/744 (17.5%).

**Table 1.** Demographics of sequenced probands.

|  | Probands (N, %) |
| --- | --- |
| Sex |  |
| Male | 406 (54.6%) |
| Female | 338 (45.4%) |
| Age* | Median, IQR |
|  | 10.2 (4 – 36) |
| Imputed ancestry** |  |
| African/African American | 22 (3.0%) |
| Ashkenazi Jewish | 33 (4.4%) |
| East Asian | 12 (1.6%) |
| European | 570 (76.6%) |
| Admixed American | 27 (3.6%) |
| Middle Eastern | 2 (0.3%) |
| South Asian | 13 (1.7%) |
| Multiple/Unassigned | 65 (8.7%) |

\* Age unknown for 116 participants
\*\*For ancestry imputation, we computed principal components (PCs) on high-quality bi-allelic autosomal SNVs in our rare disease cohorts using the gnomAD v2 method<sup>40</sup>. The sample scores from this projection are input into the gnomAD v2 random forest model and an ancestry assigned to all samples for which the probability of that ancestry is > 90%. All samples that do not meet the 90% threshold are assigned as Multiple/Unassigned.

For the 138/744 (18.5%) cases with VUS, most were in strong candidate novel disease genes (109/138, 80.0%) where ACMG/AMP criteria are not yet appropriate to be applied and the remainder were novel variants in known disease genes (29/138, 21.0%) identified that did not yet have sufficient evidence for a pathogenic classification. A list of candidate disease genes is provided and cases have been submitted to Matchmaker Exchange (Table S3).^26^

The average diagnostic yield varied significantly by phenotype category (*p*<0.0001 by chi-square test), with the highest yields seen in probands with neurodevelopmental conditions or syndromic anomalies (Figure 2). Our solve rate did not significantly differ by imputed ancestry (Table S3) either when considered across multiple groups or when all European were compared to non-European ancestries, though this may be due to the small sample size as the solve rates were lower for African/African American, Admixed American, and East Asian (22%, 15%, 17%) as compared to European (43%).

**Figure 1.**
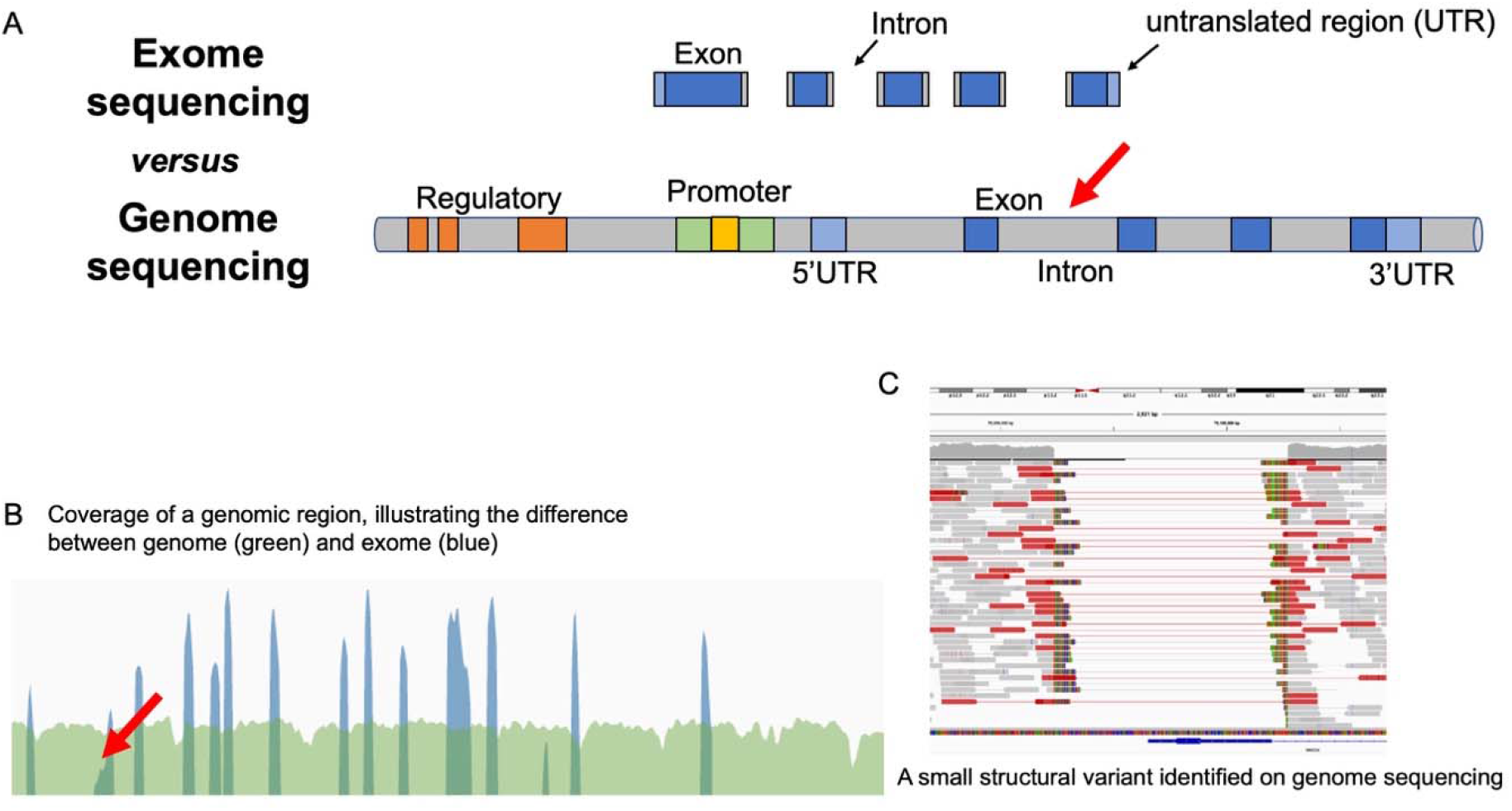
Variants assessed as requiring genome sequencing over exome sequencing to be identified within our cohort. These include: (A) deep intronic variants (arrow) unlikely to be reliably detected by typical ES methodologies (greater than 20 base pairs upstream/downstream from the beginning or end of an exon), (B) indels or SNVs that were missed on prior ES due to poor coverage of the region (arrow), tandem repeat expansions (TREs), and some SVs such as copy-neutral inversions, (C) small (∼50-2000 base pair) and/or largely intronic CNVs, or complex events involving more than one type of SV.

**Figure 2.**
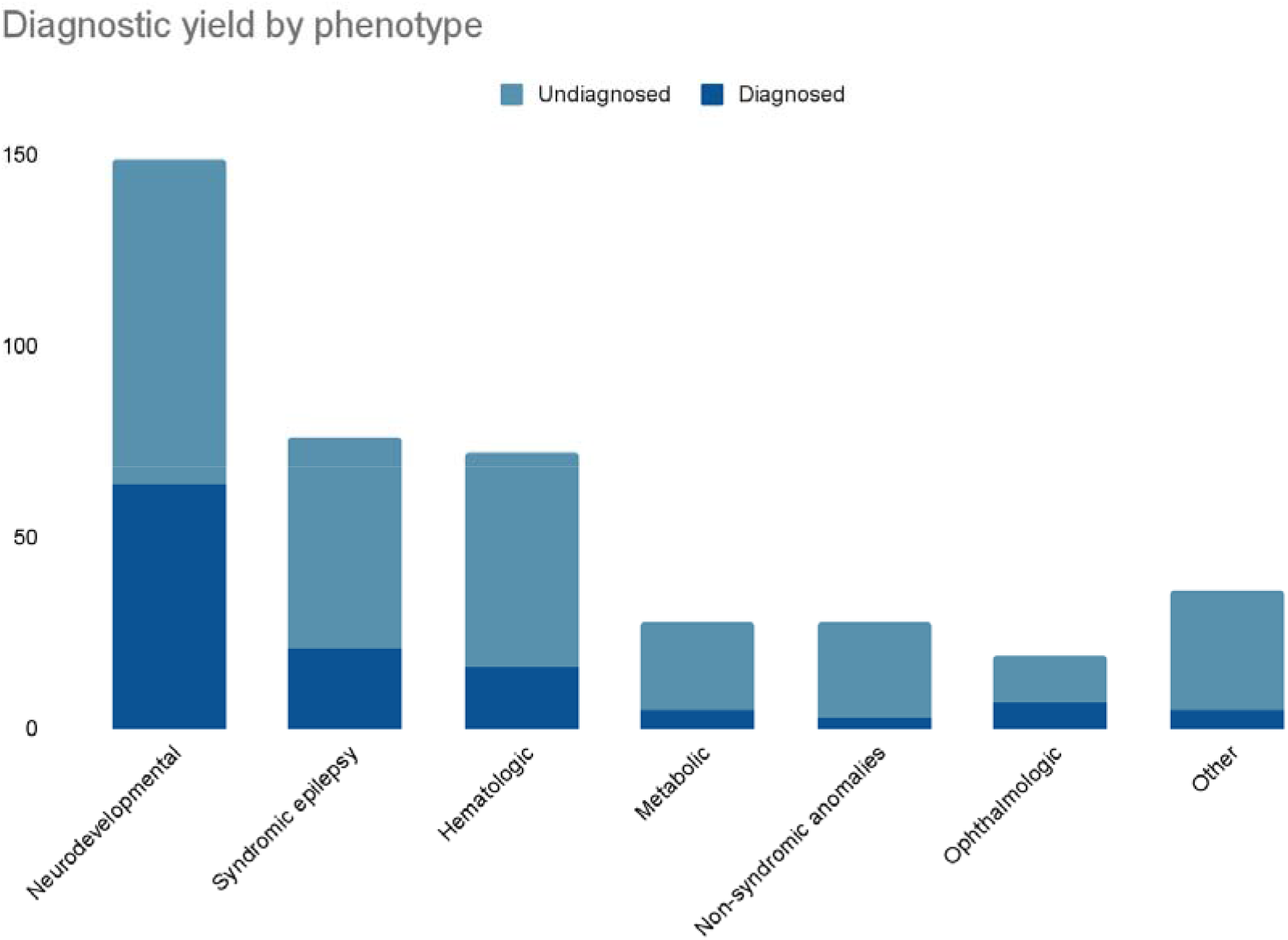
Diagnostic yield by phenotype category. Probands were categorized by most prominent phenotypic features.

### Solves requiring GS

We determined that 62 of our 218 solved/likely solved cases required GS to detect the causal variant, comprising 28.3% of the diagnosed cases and 8.3% of the entire cohort (Figure 3). Of these 62 cases, 55 had prior negative ES, and two others had prior targeted sequencing of the causal gene (*ADA, DMD*). The remaining five cases were empirically determined to require GS as they involved a deep intronic variant, a complex SV and three TREs unlikely to be identified via ES pipelines.

**Figure 3.**
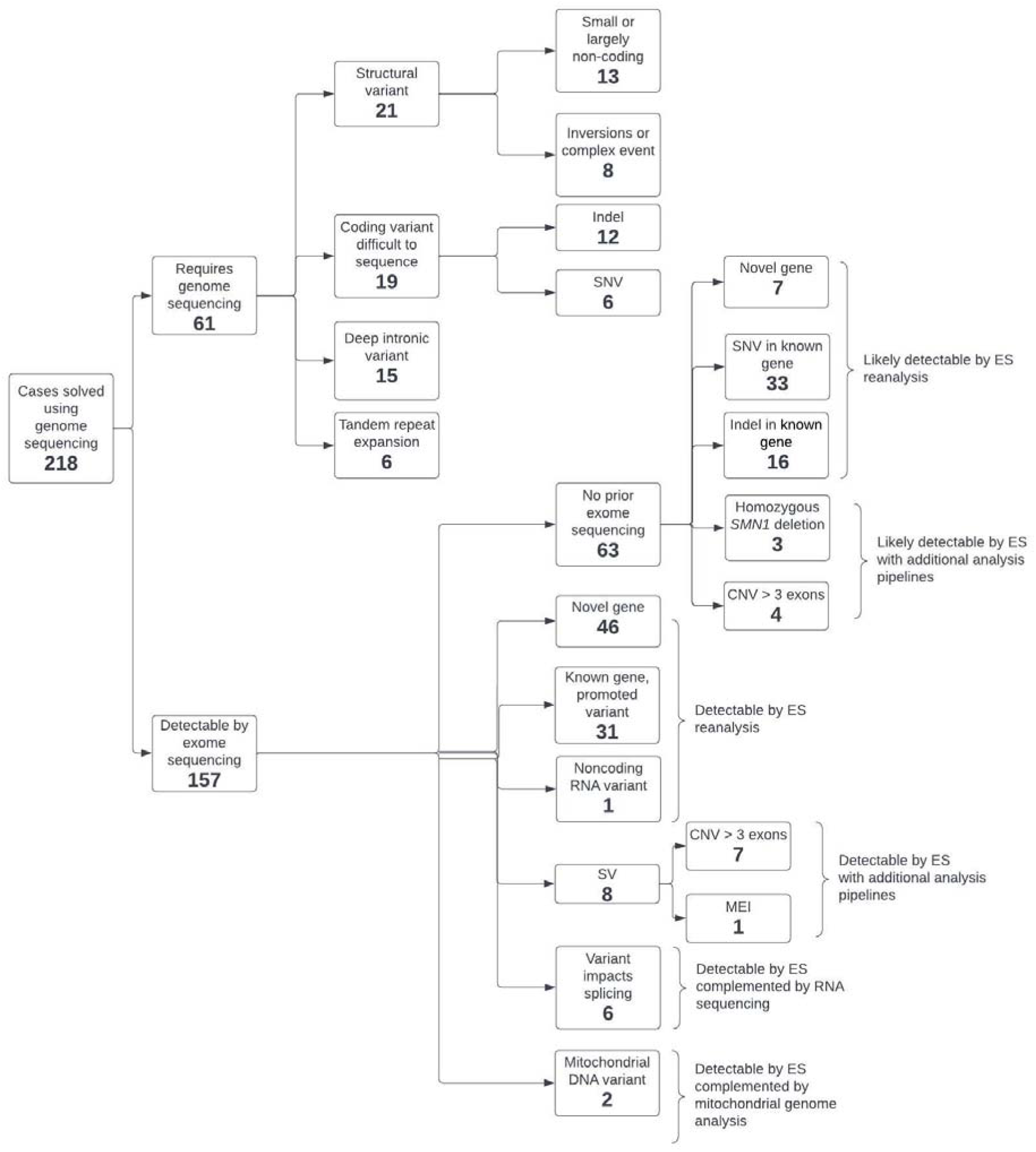
Classification of solved cases. Of all 218 cases solved/likely solved by GS, the types of variants requiring GS (61) to identify are displayed. Twenty-one (34%) were SVs, including 11 deletions (2 non-coding regions), 2 duplications, 4 inversions, 3 complex del/dup events, and one mobile element insertion (MEI) in a non-coding region. Additionally, there were 6 tandem repeat expansions identified. Nineteen cases (31%) requiring GS involved coding variants missed on prior ES due to poor coverage of the region, either resulting in no variant call at that site or a poor-quality variant that was filtered out during the analysis process. Most of these (13/19) were indels (small insertion/deletions usually of 10-15 bp in size) of which 8/13 were deletions and 5/13 were involved indels of different types. Fifteen (25%) of the diagnoses requiring GS involved deep intronic non-coding variants (complemented by RNA sequencing). Three cases had variants that were missed for more than one reason (SVs in non-coding regions for two and a deep intronic variant in a novel gene in one). Of the cases solved by variants that were detectable by ES, most (93/156, 59.6%) had prior ES that missed the variant, whereas the remaining 63 did not have prior ES but were solved via variants expected to be detectable by current ES methodologies. The three cases of *SMN1* homozygous deletions responsible for spinal muscular atrophy (SMA) were identified in RGP probands (two were concurrently identified via clinical genetic testing, one was an adult who had been clinically diagnosed with SMA of unknown type but never received testing). Two diagnostic mitochondrial DNA (mtDNA) variants were identified by GS and missed by prior ES, although these variants were also identified by mitochondrial genome sequencing.

### Diagnostic yield of GS detectable by ES

Most cases solved via GS previously had non-diagnostic ES (148/218, 67.9%) (Figure 4), confirming that these variants were overlooked previously. Overall, 94/148 (63.5%) exome-negative cases later solved by GS could have been found by exome reanalysis, most commonly because the diagnosis involved a recent novel gene-disease discovery (46/94, 48.9%), a variant in a known gene that was re-interpreted as disease causing over time (31/94, 33.0%), or analysis of variants in noncoding disease-associated transcripts (1/94, 1.1%).

**Figure 4.**
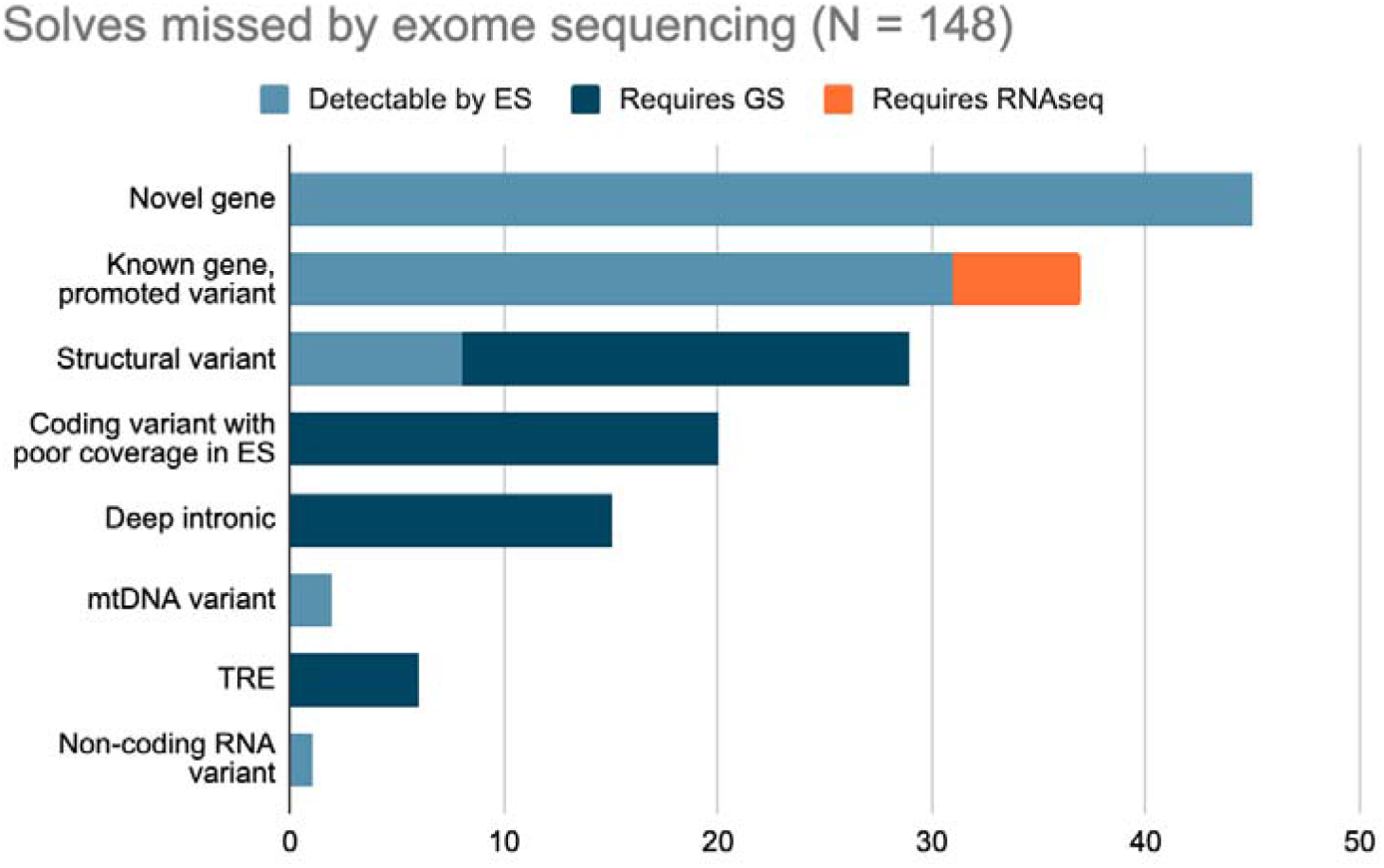
Reasons cases were not solved by prior exome sequencing.

Expanding the methodologies for variant calling applied to ES is predicted to uplift diagnosis by 3.2% (24/744) overall for the cohort. If CNV calling (8), mobile element insertion calling (1),^27^ and mitochondrial genome variant calling (2) were performed on ES data, an additional 11/94 (11.7%) would be solved. RNA sequencing analysis helped identify or validate the impact of a variant on gene expression and/or splicing for deep intronic variants (6/94, 6.4%). For the 63 families without prior ES that could have been found by exome analysis, seven diagnoses required CNV calling (4) or specialized calling of *SMN1* deletions responsible for SMA (3).^28^ Indels were a common source of variants requiring GS, so it remains possible that some of the 16 coding indels identified in individuals without prior ES may be missed by ES. Given that this cannot be ascertained with the available data, we have not counted them as requiring GS to avoid over-reporting.

### Maximizing the yield of GS

Of the 61 solves/likely solves requiring GS, 54 (89%) were in known disease genes and most were identified by untargeted analysis either due to improved coverage of coding regions (19), or by genome-wide evaluation for rare SVs (21) or known pathogenic TREs (6). Deep intronic variants (15) were typically identified in a more targeted fashion, by evaluating genes associated with the specific phenotype, and the impact of several were validated by RNA sequencing (7/15) or other functional evaluation (2/15). Additionally, eight diagnoses involving SVs benefited from a strong clinical suspicion based upon phenotypic or biochemical data that was used to identify the underlying causal variant(s) that had been missed by standard genetic testing methodologies (*FBN1, ADA, QDPR, NIPBL, RPGRIP1, DMD, RSP19, EDA*). ^29,30^ In these cases, a clinical diagnosis suggested a particular molecular genetic diagnosis - often narrowing the differential to a single gene - though the diagnosis eluded either ES or more targeted testing.

Only seven solved cases requiring GS were in novel disease genes, reflecting the challenges in prioritizing and validating variants beyond ES for genes not currently associated with human disease and the power of GS analyzed via research consortia. These diagnoses include: i) a homozygous deep intronic variant in a novel mitochondrial disease gene, *NDUFB10*, ultimately solved via a ‘multi-omic’ approach;^31^ ii) a deep intronic variant in *RPL17*; iii) a homozygous splice-impacting variant in *CYS1* missed on ES due to poor coverage of a GC-rich region; ^32^ iv) a structural long non-coding RNA variant in *CHASERR*; v) a missense variant in *PNPLA7* that was missed due to poor coverage on ES present in trans with a splice-impacting variant; vi) a single exon deletion in the loss-of-function-constrained *ZFHX3* gene; vii) and a homozygous 16 kb deletion in *WBP4* identified on GS complemented by RNA sequencing.^33^ The latter four diagnoses were aided by identification of other affected cases via the Matchmaker Exchange.^34^

## Discussion

We describe a large cohort of individuals with rare disease for whom a plausible diagnosis was found using GS, often after being missed by prior testing approaches. These cases illustrate several types of pathogenic variation that may be missed by ES or other standard genetic testing methodologies, particularly small or copy-neutral SVs, deep intronic variants, and TREs.

Importantly, these cases were solved using short read GS, which is currently clinically available. Long read GS, which is anticipated to soon be clinically available, should also be able to identify these variants described here as well as additional findings undetectable by short read GS. For this cohort, in order to achieve the overall solve rate of 29.3% after prior negative testing, 13.3% of families needed ES reanalysis, 2.3% needed additional methodologies (calling of CNV, mitochondrial genome variants, mobile element insertions, and *SMN1* deletions on ES data), and 8.3% of families truly required sequencing of the genome.

Most of our solved and likely solved cases requiring GS (47/61, 77%) were identified in an untargeted fashion, by systematically applying analytic tools across the entire cohort. As most diagnoses identified in the cohort could have been found using ES, those that truly required GS offer important insight into optimal clinical application of GS for rare disease diagnosis. Our analysis for SVs was particularly high-yield at identifying variants missed by prior testing, as 35 solves involved SVs, most (20/35) of which required GS to identify. The deep intronic variants described in this cohort were identified by a targeted approach, narrowed down to a single gene or list of genes by phenotyping or RNA sequencing. This reflects not only the challenges in variant prioritization related to the multitude of SVs, deep intronic, or non-coding variants identified by GS data, but also suggests that additional diagnoses may be found in the future by continued reanalysis of GS data from unsolved cases using regularly updated candidate gene lists. These diagnoses also demonstrate the value of careful phenotyping in the evaluation of genetic testing results in addition to prioritization of genetic loci for evaluation by GS via transcriptome analysis (RNA sequencing).^35^ In particular, a phenotypically-driven or otherwise targeted approach may mitigate some of the analytic burden of GS analysis, as a deep analysis of a limited number of genetic loci is more feasible than a search for SNVs and SVs across the entire genome. We have previously demonstrated the success of this technique in an individual with early-onset Marfan syndrome, in which a structural variant (deletion) was identified using GS that had been missed by multiple prior approaches, including targeted deletion/duplication analysis via multiplex ligation-dependent probe amplification (MLPA) in addition to ES.^30^ Critical to this approach is the ability to accurately phenotype the patient in order to guide the genomic evaluation^36^, and, indeed, the diagnostic yield within our cohort was greatly augmented by the expertise of the investigators providing cohorts for collaborative analysis.

Prior evaluations of the diagnostic yield of GS when compared to ES have demonstrated either a similar^13^ or only mildly increased yield^11^, with few diagnoses identified that would not be detectable by ES alone - usually SVs.^4,12–14,37,38^ In particular, one prior report of 108 cases of GS after non-diagnostic ES identified an incremental yield of 7%, but only 3% for cases requiring GS for diagnosis.^11^ A more recent study of GS as a first-line modality for rare disease diagnosis via identified an incremental yield of 37.5% in those who had prior ES, though notably only 16 individuals (1.5% of the larger cohort) had ES prior to GS, making it difficult to directly compare to our data given the small numbers^39^. Furthermore, while 14% of all diagnoses were described by the investigators as requiring GS for detection, this included all SVs identified in the study, even though some may be callable on ES data, which is distinct from our approach.^39^ Our larger proportion of diagnoses requiring GS for detection (28.4% of the solved cohort) compared to this and prior studies also likely reflects our increased detection of SVs and variants outside of coding regions, as many prior analyses of GS yield have focused primarily on coding variants^5,13,14^, as well the case selection in our cohort, with families with a high probability of monogenic disease (often with prescreening by gene panel or prior ES) selected for sequencing via the Broad CMG.

Overall, our results support the use of GS as a first-line test for rare disease diagnosis due to the ability to detect multiple types of disease-causing variation, replacing both ES and chromosomal microarray in a single test, and superior ability to detect all classes of variation (with the exception of somatic mosaic variants, for which the lower mean coverage of GS reduces sensitivity). However, routinely applying GS after ES or other non-diagnostic testing is still not likely to be the most efficient strategy, reflected in our relatively low incremental yield after non-diagnostic ES. While sequencing, analysis, and storage costs for short read GS and variation in payor reimbursement may present limitations to its routine application for genetic diagnosis, we anticipate these barriers to be overcome in the future, particularly as sequencing costs continue to fall. For people who remain undiagnosed after clinical ES, our findings support a reasonable diagnostic potential from GS, especially in the case where there is a clinical diagnosis directing focused attention in the GS analysis. Transcriptome analysis, which was utilized for a minority of families here, has a complementary role in GS, and its implementation in clinical testing laboratories and is likely to further improve diagnosis rates. Altogether, our findings provide important insight into the present diagnostic utility of GS and the role it may play in the future.

## Data Availability

All data produced are available online at

https://www.ncbi.nlm.nih.gov/projects/gap/cgi-bin/study.cgi?study_id=phs001272.v1.p1

## Acknowledgements

We thank the many families who participate in the Rare Genomes Project or via our Broad CMG collaborators research studies for sharing their samples and medical data.

