## Supplement for "Unique Capabilities of Genome Sequencing for Rare Disease Diagnosis"

**Contents:**

**Methods**……………………………………………………………………………………...2

**Sequencing**…………………………………………………………………………..2

**Analysis**……….………………………………………………………………………..3

**Variant classification**……………………………...…………………………………..5

**References**…………………………...…………………………...…...……………………..5

**Table S1 Standard searches used by the Broad team for ES/GS analysis**…………...…...…...9

**Figure S1 Classification of variants**..……………………….……………………...…...…...10

**Table S2. Diagnoses**..…………………………………………...……………...…...…...11

**Table S3. Candidates**..……………………………………….…………………...…...…...12

**Table S4. Diagnostic yield by imputed ancestry**..………………………………...…...…...15

### Sequencing Methods

Genome sequencing (GS) was performed by the Genomics Platform at the Broad Institute of MIT and Harvard. PCR-free preparation of sample DNA (350 ng input at >2 ng/ul) is accomplished using Illumina HiSeq X Ten v2 chemistry. Libraries are sequenced to a mean target coverage of 30x. GS data was processed through a pipeline based on Picard, using base quality score recalibration and local realignment at known indels. The BWA aligner was used for mapping reads to the human genome build 38 (GRCh38). Single nucleotide variants and insertions/deletions (indels) were jointly called across all samples using Genome Analysis Toolkit (GATK) HaplotypeCaller package version 4.0. Default filters were applied to SNV and indel calls using the GATK Variant Quality Score Recalibration (VQSR) approach. Annotation was performed using Variant Effect Predictor (VEP). GATK-SV[1] was used to detect structural variants (SVs), which were annotated with the GATK SVAnnotate tool. Mitochondrial DNA (mtDNA) single nucleotide and small indel variants were called from GS data using the gnomAD-mitochondria pipeline[2] and large mtDNA deletions were called by MitoSAlt[3]. ExpansionHunter v5 was used to genotype known disease-associated tandem repeat expansions (TREs).[4] Lastly, the variant call set was uploaded to *seqr* for collaborative analysis between the CMG and investigator or for analysis by the RGP team.[5]

ES was performed prior to GS for many of these cases through a variety of clinical diagnostic laboratories or by the Genomics Platform at the Broad Institute. For these cases, libraries from DNA samples (>250 ng of DNA, at >2 ng/ul) were created with an Illumina Nextera exome capture (38 Mb target) and sequenced (150 bp paired reads) to cover >80% of targets at 20x and a mean target coverage of 80x until January 2019 and thereafter using a Twist exome capture (~38 Mb target) and sequenced (150 bp reads) to cover > 90% of targets at 20x and a mean target coverage of 80x. Sample identity quality assurance checks were performed on each sample. The ES data was de-multiplexed and each sample's sequence data were aggregated into a single Picard BAM file. ES data was subsequently processed similar to GS data as previously described.

**Analysis**

The Broad CMG analysis team has developed four standard searches that are applied for each family (Table S1). We prioritize variants for further study that have high pathogenicity scores using common *in silico* predictors (e.g. REVEL[6], CADD[7], SIFT[8], PolyPhen-2[9], MutationTaster[10], MPC[11]), occur at highly conserved residues as determined by manual review on the UCSC genome browser and evaluating the Genomic Evolutionary Rate Profiling (GERP) score[12]. We visually inspect the read data using the Integrated Genomics Viewer (IGV) for our candidate variants to ensure they are not sequencing artifacts. Top candidate variants are typically confirmed by an orthogonal method such as Sanger sequencing.

To detect SVs, we initially utilized multiple SV-calling tools including Manta[13], DELLY[14], and Smoove (https://github.com/brentp/smoove), and more recently applied GATK-SV: an ensemble SV detection tool that discovers, genotypes, and resolves the diverse classes of SVs that can be captured from GS data, including balanced and unbalanced CNVs, inversions, insertions, translocations, and a spectrum of complex SVs. Briefly, GATK-SV maximizes sensitivity by harmonizing five algorithms, then adjudicating and re-genotyping SVs from raw read evidence[1]. GATK-SV considers all SV evidence available from GS, including discordant paired-end (PE) or split reads (SR) crossing a breakpoint, and normalized read-depth (RD) or B-allele frequencies. Each CRAM file is processed with five algorithms, which currently include two PE/SR algorithms (Manta[13], Wham[15]), two RD algorithms (cnMOPS[16] and GATK-gCNV[17]), and a mobile element algorithm, MELT[18]. GATK-SV is publicly available on GitHub (https://github.com/broadinstitute/gatk-sv). For CNV analysis, we also apply germline Copy Number Variant caller (gCNV), a coverage-based CNV detection method that normalizes coverage across the exome by adjusting for systematic bias and uses a probabilistic framework to infer copy number from the normalized coverage. We manually evaluate the CNV data, filtering out low-quality calls and inherited variants (based on family history) and focus our analysis on CNVs overlapping protein-coding genes. For cases with a strong phenotype pointing to a particular gene or genes as the likely candidate, we may also manually search for SVs by visually-inspecting the reads across the gene in question (using the Integrated Genomics Viewer[19]).

To evaluate for tandem repeat expansions (TREs), we run ExpansionHunter v5[20] on GS samples to genotype 60 known disease-associated repeat loci. The locus specifications we use are publicly available on github (https://github.com/broadinstitute/str-analysis) and represent the same list of loci for which population frequencies are available in the gnomAD browser (https://gnomad.broadinstitute.org/short-tandem-repeats?dataset=gnomad_r3). We also run REViewer[21] to generate read visualizations. Then, to identify candidate pathogenic expansions, we evaluate individuals with the most-expanded genotypes for each locus, comparing them to the pathogenic threshold and population frequencies for this locus in gnomAD. We also evaluate genotype qualities based on reviewing read visualizations.

To evaluate mtDNA SNVs and indels, we run the mitochondria mode of Mutect2 followed by the gnomAD-mitochondria pipeline[2]. Then, to identify candidate variants, we search for "confirmed" variants listed in MITOMAP[22] and P/LP variants listed in ClinVar. We finally review all mtDNA variants of uncertain significance (VUS) reported in ClinVar and/or with “reported” status in MITOMAP, in addition to unreported variants with pathogenic *in silico* prediction based on mtDNA-specifications of the ACMG/AMP guidelines (APOGEE $\geq$ 0.5 for missense variants; MitoTIP > 12.66 plus HmtVar $\geq$ 0.35 for tRNA variants), that are absent at high heteroplasmy level ($\geq$80%) or homoplasmy ($\geq$ 95%) in reference databases (gnomAD v3 and HelixMTdb).

**Variant classification**

In order to systematically assess the pathogenicity of the structural variants that we identified, the American College of Medical Genetics and Genomics (ACMG) and the Clinical Genome Resource (ClinGen) standards for classification and reporting of constitutional copy-number variants were applied[23]. Variants in novel gene-disease relationships are classified as VUS until the gene-disease relationship has at least moderate evidence supporting it.  CNV associated with disorders that follow an autosomal recessive or X-linked mode of inheritance are not addressed in these standards and required additional consideration; the classification criteria were modified to optimally capture evidence for pathogenicity for the range of variants that we identified. Relative proportions of VUS to pathogenic/likely pathogenic variants are presented in Figure S1.

**Table S1: Standard searches used by the Broad team for ES/GS analysis**

| Search | Variant annotations | Variant frequency (Broad callset, gnomAD[24], gnomAD SV[1]) | Variant call quality | Annotation overrides (SpliceAI score[25]) |
|---|---|---|---|---|
| Dominant/*de novo* restrictive | Coding variants, essential and extended splice site, LOF SVs | 0.001 gnomAD<br><br>0.01 callset | Pass VQSR<br>GQ 40<br>AB 20 | ClinVar LP/P<br><br>SpliceAI >0.2 |
| Recessive restrictive | Coding variants, essential and extended splice site, LOF SVs | 0.01 gnomAD<br><br>0.03 callset | Pass VQSR<br>GQ 40<br>AB 20 | ClinVar LP/P<br><br>SpliceAI >0.2 |
| Dominant/*de novo* permissive | Coding variants, synonymous, splice, 5/3'UTR, Non-coding exons, TFBS, regulatory region, LOF/intronic/UTR/promoter SVs | 0.001 gnomAD<br><br>0.01 callset | GQ 40<br>AB 10 | ClinVar LP/P/VUS<br><br>SpliceAI >0.1 |
| Recessive permissive | Coding variants, synonymous, splice, 5/3'UTR, Non-coding exons, TFBS, regulatory region, LOF/intronic/UTR/promoter SVs | 0.01 gnomAD<br><br>0.03 callset | GQ 40<br>AB 10 | ClinVar LP/P/VUS<br><br>SpliceAI >0.1 |

Legend: LOF: Loss of function; VQSR: Variant Quality Score Recalibration; GQ: Genotype quality; AB: allele balance; LP/P: Likely pathogenic/pathogenic; UTR: Untranslated region; TFBS: transcription factor binding site; VUS: Variant of uncertain significance

**Figure S1. Classification of variants.** Pathogenicity of 284 variants in 218 families solved via GS, classified as per the ACMG/AMP/ClinGen standards.

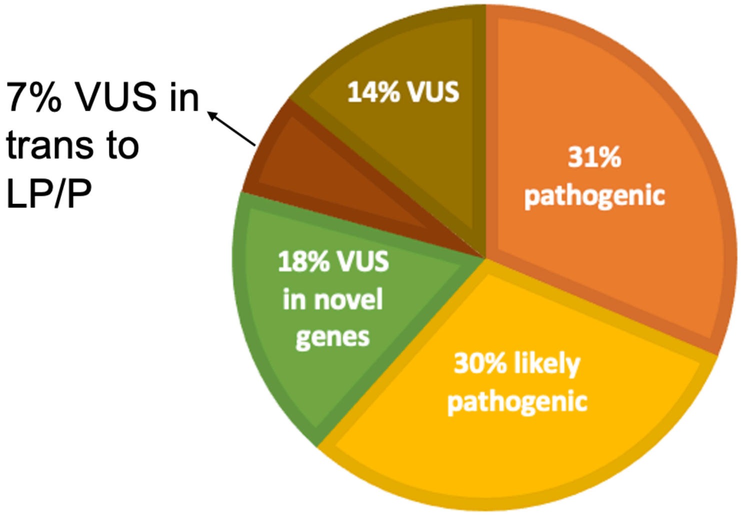

**Table S2. Diagnoses**
See separate spreadsheet

**Table S3. Candidates.** Candidate novel disease genes identified in this cohort.

| **ID** | **Gene** |
|---|---|
| UWA_LAI963 | *ABCD3* |
| HK115 | *ACSL5* |
| SCO_PED096 | *ADGRE3* |
| BON_B17-59 | *AFAP1L1* |
| SOU_FAM00008 | *AGMAT* |
| HK103 | *ANO1* |
| HK081 | *ANO2* |
| RGP_696 | *ARFGEF3* |
| CHU_05 | *ARHGAP6* |
| RGP_245 | *BAZ1B* |
| RGP_12 | *BLOC1S1* |
| RGP_655 | *BOD1* |
| CHU_23 | *C10orf71* |
| RGP_572 | *CACNA2D3* |
| RGP_658 | *CAMK1D* |
| RGP_284 | *CAMK4* |
| HK060 | *CBX8* |
| HK017 | *CDK11B* |
| RGP_735 | *CDK16* |
| IK | *CDK5RAP3* |
| RGP_1374 | *CEP192* |
| BEG_0761 | *CFAP46* |
| RGP_868 | *CFAP54* |
| CMG_Laing_Ravencroft_WGS | *COL5A3* |
| RGP_1333 | *DIPK2B* |
| RGP_45 | *DNAH17* |
| RGP_1149 | *EBF2* |
| 49 | *ELK1* |
| UWA_LAI1646 | *EP400* |
| RGP_726 | *EPHA6* |
| RGP_54 | *ERICH3* |
| RGP_1268 | *ETV1* |
| 827 | *FAM193A* |
| RGP_1180 | *FBXO42* |
| RGP_375 | *FGF7* |
| HK085 | *FLYWCH1* |
| RGP_1150 | *FRG2* |

| | |
|---|---|
| RGP_86 | *FRMPD3* |
| RGP_589 | *FURIN* |
| RGP_1129 | *GFPT1* |
| RGP_119 | *GRM4* |
| HK028 | *GTF2A1* |
| HK032 | *HAPLN2* |
| HK044 | *HEATR1* |
| HK075 | *HELZ* |
| RGP_673 | *HNRNPL* |
| VCGS_FAM148 | *HOXC8* |
| RGP_1016 | *INTS6L* |
| CHU_04 | *ISLR2* |
| RGP_1479 | *KCNH8* |
| RGP_95 | *KDM4A* |
| RGP_1392 | *KDM8* |
| HK080 | *KIAA0408* |
| RGP_1526 | *KLHL13* |
| FAM39 | *LATS2* |
| RGP_1498 | *LBX1* |
| RGP_289 | *MACO1* |
| RGP_105 | *MARCH5* |
| RGP_20 | *MAU2* |
| HK104 | *MCRS1* |
| RGP_674 | *MRPL54* |
| RGP_1175 | *MYH1* |
| BON_B18-54 | *MYO7B* |
| RGP_526 | *NCOR1* |
| RGP_1425 | *NCOR2* |
| RGP_495 | *NELL2* |
| RGP_314 | *OSBPL9, SYNRG* |
| RGP_1138 | *PACSIN3* |
| RGP_329 | *PKP4* |
| RGP_1193 | *PPP1R12C* |
| RGP_232 | *PRICKLE3, GNA13* |
| RGP_1504 | *PRPF4B* |
| VCGS_FAM52 | *PRPS2* |
| RGP_853 | *PTPRG* |
| RGP_53 | *RAB33A* |
| MAN_1601 | *RCOR2* |

| | |
|---|---|
| RGP_230 | *RHPN2* |
| RGP_469 | *RIMS1* |
| 235 | *RPL37A* |
| GI | *SARNP* |
| RGP_402 | *SCRIB* |
| RGP_522 | *SCTR* |
| HK035 | *SH3GL1* |
| 1024 | *SHCBP1* |
| HK072 | *SMG6* |
| RGP_1099 | *SMYD1* |
| FAM61 | *SNED1* |
| RGP_682 | *SRGAP2* |
| RGP_731 | *SRRT* |
| RGP_5 | *SSBP3* |
| RGP_135 | *SSH1* |
| RGP_918 | *SYNM* |
| CHU_01 | *TBC1D22A* |
| RGP_431 | *THAP12* |
| RGP_951 | *THBS2* |
| RGP_677 | *TLK1* |
| RGP_123 | *TPPP* |
| FAM29 | *TPR* |
| VCGS_FAM147 | *TRABD2B* |
| 966 | *TTC28* |
| RGP_452 | *TXLNG* |
| RGP_1101 | *UNC13B* |
| VCGS_FAM2 | *UNC5C* |
| RGP_1125 | *VPS37D* |
| FAM1 | *WWP1* |
| RGP_504 | *ZBTB1* |
| RGP_763 | *ZC3H11A* |

**Table S4. Diagnostic yield by imputed ancestry**

| **Ancestry Category** | **Total**<br><br>**(N, % of cohort)** | **Diagnosed**<br><br>**(N, % of subgroup)** |
|---|---|---|
| African/African American | 22 (3.0%) | 5 (22%) |
| Ashkenazi Jewish | 33 (4.4%) | 20 (61%) |
| East Asian | 12 (1.6%) | 2 (17%) |
| European | 570 (76.6%) | 170 (43%) |
| Latino/Admixed American | 27 (3.6%) | 4 (15%) |
| Middle Eastern | 2 (0.3%) | 1 (50%) |
| South Asian | 13 (1.7%) | 4 (31%) |
| Multiple/Unknown | 65 (8.7%) | 19 (29%) |
